# Sex difference in incidence of bipolar and anxiety disorders: findings from the Global Burden of Disease Study 2021

**DOI:** 10.1101/2024.10.27.24316200

**Authors:** Xingke Zhu, Qing Lv

**Author notes:** **Corresponding author:** Xingke Zhu; Qing Lv. **Author contributions:** XZ and QL had full access to all the data in the study and will take responsibility for the integrity of the data, the accuracy of the data analysis and study concept and design. XZ contributed to the acquisition, analysis and interpretation of data. XZ and QL contributed to drafting of the manuscript. XZ contributed to the critical revision of the manuscript and supervision of the study. All authors read and approved the final manuscript. **Funding:** This research did not receive any specific grant from funding agencies in the public, commercial, or not-for-profit sectors. **Compliance with Ethics Requirements:** For Global Burden of Disease study, a waiver of informed consent was reviewed and approved by the Institutional Review Board of the University of Washington. All the information about ethical standards is available through the official website (https://www.healthdata.org/research-analysis/gbd). **Ethics approval and consent to participate:** Not applicable.

## Abstract

**Background:** Bipolar disorder and anxiety disorders are two prominent mental disorders that represent a significant global health challenge.

**Objective:** The Global Burden of Disease Study 2021 (GBD 2021) was employed to evaluate sex differences in the incidence of bipolar disorder (BD) and anxiety disorder (ANX) globally by year, age, and socioeconomic status.

**Method:** We estimated sex-specific incidence of BD and ANX from GBD 2021 globally and in 204 countries and territories from 1990 to 2021. The sociodemographic index (SDI) was used to gauge national socioeconomic development and the Health Organization (WHO) region was used as a division of regions. Differences in age-standardized incidence rates (ASRs) by sex (absolute and relative) and risk ratios (95% confidence interval) were calculated annually and by age. Annual percent change (APC) was calculated by joinpoint regression modeling and linear regression analyses were performed to explore the socioeconomic factors associated with sex differences in incidence.

**Results:** The absolute and relative sex difference in ASRs of BD showed a slight declining trend during 1990 and 2021, with absolute difference decreasing from 2.50 to 1.83, and relative difference decreasing from 1.08 to 1.06; The absolute and relative sex difference in ASRs of ANX showed an increasing trend during 1990 and 2021, with absolute difference increasing from 170.02 to 208.08, and relative difference increasing from 1.35 to 1.36. Worldwide, females had a higher risk of BD and ANX than males in 1990and 2021. The highest Risk ratios of BD and ANX were observed in the European Region in 2021.The greatest relative sex difference of BD was 1.09 in the age group of 30-34. The greatest relative difference of ANX was 1.51 in the age group of 20-24. Relative sex differences of BD and ANX were significantly and positively correlated with SDI (BD, standardized β = 0.27 (95% CI, 0.22 to 0.33), *P* < 0.001; ANX, standardized β = 0.80 (95% CI, 0.47 to 1.14), *P* < 0.001).

**Conclusions:** Sex difference in the incidence of anxiety disorders and bipolar disorder have persisted worldwide over the past several decades, and the rates have consistently been higher among females than males. The sex difference in the global incidence of bipolar disorder has shown a slight improvement, but that in the global incidence of anxiety disorders has not been effectively mitigated. The sex difference is even more pronounced at younger ages and in more developed nations. The findings emphasize the significance of sex-specific health policies to reduce sex differences in the incidence of bipolar and anxiety disorders.

## Introduction

Bipolar disorder and anxiety disorders are two prominent mental health conditions that impact millions of individuals across the globe [1, 2]. Anxiety disorders involve persistent and excessive worry, fear, or anxiety that interferes with an individual’s ability to function effectively. Worldwide, 301.4 million people suffer from anxiety disorders [3]. Bipolar disorder, on the other hand, is characterized by extreme mood swings, including manic and depressive episodes, which can significantly impair daily functioning and quality of life. According to GBD 2019, anxiety disorders account for 39.5 million cases [3]. Both conditions have been shown to have substantial socio-economic and health impacts. Emotional disorders, high psychosocial morbidity, and impulsive behaviors are the characteristics of bipolar disorder patients, which also increase the risk of suicide in bipolar disorder patients [4]. According to a study on the suicidal tendencies of bipolar disorder patients, there is a significantly increased risk of attempting and dying by suicide among bipolar disorder patients [5]. Similarly, deaths from non-natural causes had the highest natural mortality rate ratio among those with anxiety disorders, and anxiety disorders were linked to a significantly greater risk of death [6]. According to the total disability-adjusted life years (DALYs) attributed to mental disorders globally in 2019, anxiety disorders and bipolar disorder ranked second (22.9%) and fifth (6.8%), respectively [3].

Sex differences in the burden of mental health disorders have been a topic of significant research interest. Sex differences in the burden of BD and ANX have been an important issue of global public concern, although psychotherapy and medication are effective treatments for them. Epidemiologic and clinical research indicates that is about two times higher amongst female adolescents as amongst male adolescents, and among bipolar disorder patients, suicide attempts were 35% more common in women than in men [7, 8]. Anxiety problems also vary by gender. Women have persistently higher prevalence rates of anxiety disorders, and anxiety disorders are associated with a greater sickness burden in women than in men [9]. Biological factors such as genetics and sex hormone changes, as well as psychosocial ones such as life experiences, discrimination, and social and economic circumstances, play important roles in clarifying why men and women experience bipolar and anxiety differently [10, 11]. Thus, global patterns of sex differences in BD and ANX incidence have important implications for the development of sex-specific health policies to reduce BD and ANX.

The Global Burden of Disease Study 2021 offers an updated and comprehensive dataset to explore these sex differences in the incidence of bipolar disorder and anxiety disorders [12]. The present study investigates the sex differences in the incidence of bipolar disorder and anxiety disorders using the data provided by the Global Burden of Disease Study 2021. The findings are expected to contribute valuable insights into the epidemiology of these disorders and highlight areas where gender-specific strategies could enhance mental health outcomes globally.

## Materials and methods

### Data using

The Global Burden of Disease (GBD) 2021 study estimated the incidence of bipolar disorder (BD) and anxiety disorders (ANX) for both genders across 20 age groups and 204 countries and territories from 1990 onwards. This study utilizes the Bayesian meta-regression tool DisMod-MR 2.1 to integrate various data sources (including published literature, surveillance, surveys, hospital records, and clinical data) that meet specific inclusion criteria [13]. Incidence rates are age-standardized according to the GBD reference population [14]. The study employs a technique for propagating uncertainty consistent with prior methodologies, with final estimates derived from the mean of 1000 simulations, and 95% uncertainty intervals (UIs) based on the 25th and 975th percentiles. Definitions of BD and ANX adhere to DSM-IV-TR or ICD-10 criteria [15]. Data extracted include global age-standardized incidence rates (ASRs) per 100,000 population from 1990 to 2021; global and World Health Organization (WHO) regional incident cases and ASRs for 1990 and 2021; and national ASRs for 204 countries and territories in 2021.

The sociodemographic index (SDI) is a composite measure reflecting a country’s income per capita, average years of schooling, and fertility rate for females under 25 years of age, providing a comprehensive assessment of socioeconomic development [16]. The GBD study calculated the SDI as the geometric mean of these three variables using the Human Development Index methodology [17]. Quintile cutoff values for analysis were determined based on 2021 country-level SDI estimates. The SDI values range from 0 to 1, with higher values indicating more advanced socioeconomic development. In 2021, 204 countries and territories were categorized into five SDI groups: low SDI (0 < SDI < 0.466), low-middle SDI (0.466 ≤ SDI < 0.619), middle SDI (0.619 ≤ SDI < 0.712), high-middle SDI (0.712 ≤ SDI < 0.810), and high SDI (0.810 ≤ SDI < 1) [18]. Specifically, the low SDI group included 34 countries, the low-middle SDI group 42 countries, the middle SDI group 41 countries, the high-middle SDI group 48 countries, and the high SDI group 39 countries.

### Statistical analyses

Global absolute (female minus male) and relative (female-to-male ratio) sex differences in incidence, along with risk ratios (RR) and their 95% confidence intervals (CIs), were calculated by year and age group. ASRs of BD and ANX were estimated using the Joinpoint Regression Model, a set of statistically linear models [19]. Annual percentage changes (APCs) and their 95% confidence intervals (CIs) were calculated. Age-standardized rates (ASRs) for males and females across 204 countries and territories were compared using the Mann– Whitney U test, accounting for multiple comparisons between socio-demographic index (SDI) groups [20]. Linear regression analyses assessed the relationship between absolute (female minus male) and relative (female-to-male ratio) sex differences in ASRs and SDI across these countries and territories. *P* values less than 0.05 were considered statistically significant. All statistical analyses were performed using R software (R Core Team, version 3.4.2, Vienna, Austria), Joinpoint software (version 4.9.1.0) from the Surveillance Research Program of the US National Cancer Institute. Zstats 1.0 (www.zstats.net) and GraphPad Prism 9.0 (San Diego, America).

### Ethics Statement

All the information about ethical standards is available through the official website (https://www.healthdata.org/research-analysis/gbd).

## Results

### Sex difference in incidence of BD and ANX by year

Over the past 30 years, the ASRs for BD in females and males has not changed much globally: Average Annual Percent Change (AAPC) = 0.0% (95% CI, 0.0% to 0.0%), *P* < 0.001, for females, between 1990 and 2021; AAPC=0.1% (0.1% to 0.1%), *P* < 0.001, for males, between 1990 and 2021 (Fig. 1A). However, from 1990 to 2021, ASRs of BD in both sexes increased at the global level, with ASRs being 32.61 (95% UI: 27.45 to 38.60) in 1990 and 33.06 (95% UI: 27.80 to 39.60) in 2021 for females, and being 30.11 (95% UI: 25.41 to 35.57) in 1990 and 31.22 (95% UI: 26.27 to 37.34) in 2021 for males. The absolute and relative sex difference in ASRs of BD showed a slight declining trend during 1990 and 2021, with absolute difference decreasing from 2.50 to 1.83 (Fig. 1B), and relative difference decreasing from 1.08 to 1.06 (Fig. 1C).

**Fig 1.**
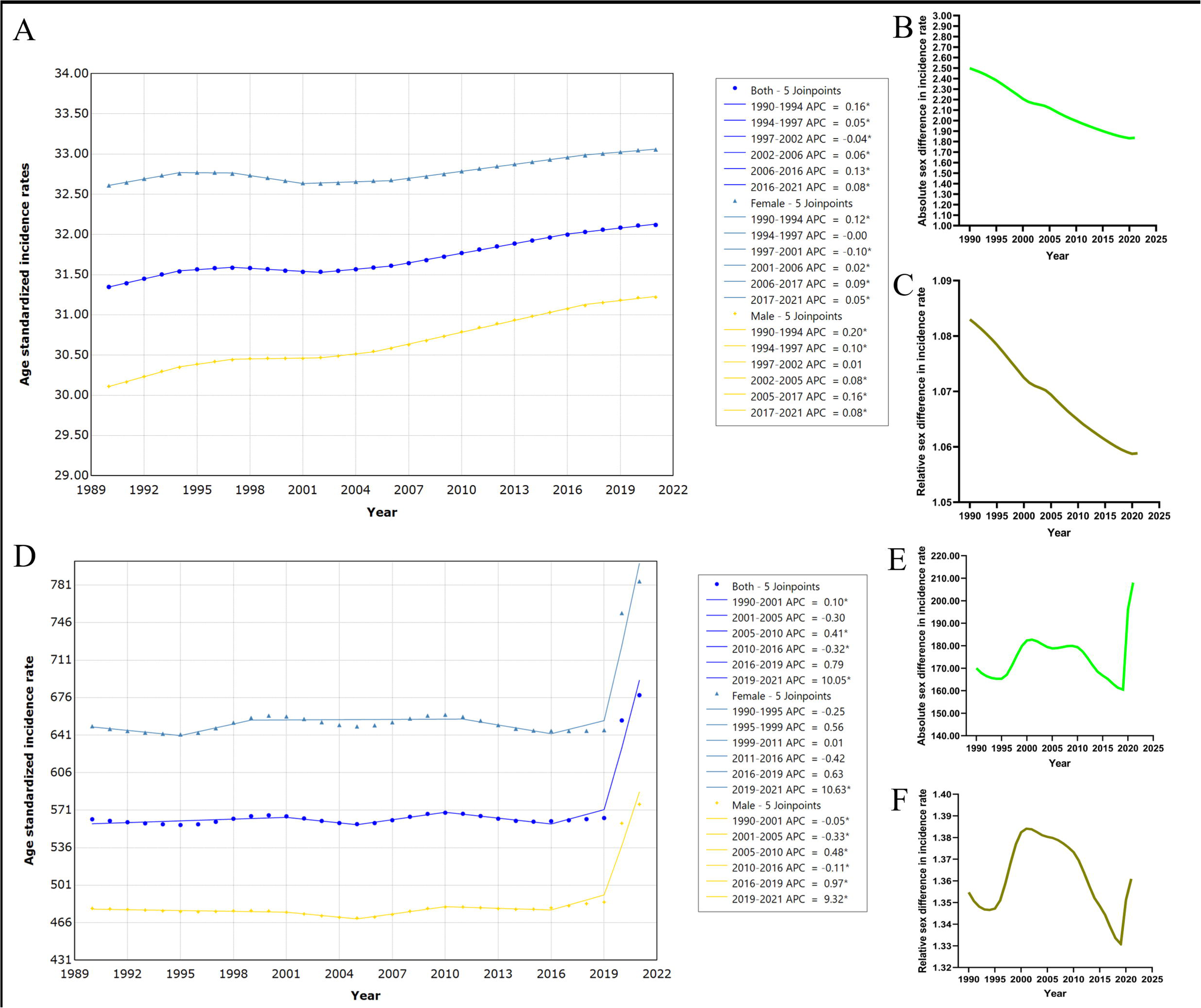
Sex difference in global incidence of BD and ANX from 1990 to 2021. (A) Age standardized incidence rates of BD. (B) Absolute (female minus male) sex difference in age standardized incidence rates of BD. (C) Relative (female to male ratio) sex difference in age standardized incidence rates of BD. (D) Age standardized incidence rates of ANX. (E) Absolute sex difference in age standardized incidence rates of ANX. (F) Relative sex difference in age standardized incidence rates of ANX.

From 1990 to 2021, the ASRs of ANX in both females and males increased globally: AAPC = 0.7% (95% CI, 0.5% to 0.9%), *P* < 0.001, for females, between 1990 and 2021; AAPC=0.6% (0.6% to 0.7%), *P* < 0.001, for males, between 1990 and 2021 (Fig. 1E). The ASR for females was 649.32 (95% UI: 537.68 to 795.75) in 1990 and 784.53 (95% UI: 649.26 to 966.02) in 2021, while the ASR for males was 479.30 (95% UI: 402.19 to 579.75) and 576.46 in 2021 (95% UI: 486.27 to 701.84). More specifically, there was a marked upward trend in the ASR of ANX from 2019 to 2021: Annual Percent Change (APC) = 10.6% (95% CI, 8.9% to 12.4%), *P* < 0.001, for females; APC = 9.3% (95% CI, 8.9% to 9.7%), *P* < 0.001, for males. The absolute and relative sex difference in ASRs of ANX showed an increasing trend during 1990 and 2021, with absolute difference increasing from 170.02 to 208.08 (Fig. 1E), and relative difference increasing from 1.35 to 1.36 (Fig. 1F).

ASRs for BD and ANX were investigated in females and males globally and in WHO regions in 1990 and 2021. Worldwide, females had a higher risk of BD that males in 1990 (RR = 1.08; 95% CI, 1.08 to 1.09) and 2021 (RR = 1.06; 95% CI, 1.06 to 1.06). The highest RRs were observed in the European Region, being 1.19 (95% CI, 1.19 to 1.20) in 1990 and 1.20 (95% CI, 1.20 to 1.921) in 2021 (Table 1). For ANX, worldwide, females also exhibited a higher risk than males in 1990 (RR = 1.35; 95% CI, 1.34 to 1.37) and 2021 (RR = 1.36; 95% CI, 1.34 to 1.38). The highest RRs were observed in the Western Pacific Region, being 1.44 (95% CI, 1.42 to 1.47) in 1990 and in the European Region, being 1.46 ((95% CI, 1.45 to 1.50) in 2021 (Table 2).

**Table 1.** The global and WHO regional incidence of bipolar disorder for both sexes in 1990 and 2021.

| Year and region | Female |  |  | Male |  |  | Risk ratio |  |  |
| --- | --- | --- | --- | --- | --- | --- | --- | --- | --- |
|  | Incidence rates | Upper | Lower | Incidence rates | Upper | Lower | Mean | Upper | Lower |
| 1990 |  |  |  |  |  |  |  |  |  |
| Global | 32.61 | 38.60 | 27.45 | 30.11 | 35.57 | 25.41 | 1.08 | 1.09 | 1.08 |
| Eastern Mediterranean Region | 44.56 | 54.49 | 36.44 | 40.96 | 49.70 | 33.34 | 1.09 | 1.10 | 1.09 |
| European Region | 47.08 | 56.63 | 39.28 | 39.41 | 47.12 | 32.75 | 1.19 | 1.20 | 1.19 |
| Region of the Americas | 56.85 | 66.28 | 49.05 | 52.19 | 60.43 | 45.11 | 1.09 | 1.10 | 1.09 |
| South-East Asia Region | 23.10 | 27.43 | 19.35 | 25.04 | 29.78 | 21.02 | 0.92 | 0.92 | 0.92 |
| Western Pacific Region | 17.66 | 20.81 | 14.91 | 16.21 | 19.02 | 13.73 | 1.09 | 1.09 | 1.09 |
| African Region | 37.82 | 46.22 | 30.91 | 36.50 | 44.42 | 29.82 | 1.04 | 1.04 | 1.04 |
| 2021 |  |  |  |  |  |  |  |  |  |
| Global | 33.06 | 39.60 | 27.80 | 31.22 | 37.34 | 26.27 | 1.06 | 1.06 | 1.06 |
| Eastern Mediterranean Region | 43.43 | 53.06 | 35.41 | 40.32 | 48.91 | 32.68 | 1.08 | 1.08 | 1.08 |
| European Region | 48.44 | 58.29 | 40.24 | 40.45 | 48.36 | 33.61 | 1.20 | 1.21 | 1.20 |
| Region of the Americas | 57.07 | 66.57 | 49.12 | 52.57 | 61.29 | 45.34 | 1.09 | 1.09 | 1.08 |
| South-East Asia Region | 23.07 | 27.39 | 19.38 | 25.14 | 29.83 | 21.14 | 0.92 | 0.92 | 0.92 |
| Western Pacific Region | 17.85 | 21.17 | 15.10 | 16.57 | 19.59 | 14.00 | 1.08 | 1.08 | 1.08 |
| African Region | 37.43 | 45.68 | 30.59 | 36.24 | 44.08 | 29.59 | 1.03 | 1.04 | 1.03 |

WHO: World Health Organization. The confidence interval is 95%.
Incidence rates: Age-standardized incidence rates.

**Table 2.** The global and WHO regional incidence of anxiety disorder for both sexes in 1990 and 2021.

| Year and region | Female |  |  | Male |  |  | Risk ratio |  |  |
| --- | --- | --- | --- | --- | --- | --- | --- | --- | --- |
|  | Incidence rates | Upper | Lower | Incidence rates | Upper | Lower | Mean | Upper | Lower |
| 1990 |  |  |  |  |  |  |  |  |  |
| Global | 649.32 | 795.75 | 537.68 | 479.30 | 579.75 | 402.19 | 1.35 | 1.37 | 1.34 |
| Eastern Mediterranean Region | 752.87 | 946.05 | 615.88 | 605.97 | 733.70 | 504.65 | 1.24 | 1.29 | 1.22 |
| European Region | 733.96 | 913.49 | 605.57 | 516.01 | 627.22 | 431.41 | 1.42 | 1.46 | 1.40 |
| Region of the Americas | 807.89 | 1006.83 | 664.27 | 602.69 | 729.42 | 503.77 | 1.34 | 1.38 | 1.32 |
| South-East Asia Region | 562.46 | 688.52 | 460.86 | 432.02 | 519.03 | 362.05 | 1.30 | 1.33 | 1.27 |
| Western Pacific Region | 620.06 | 750.29 | 516.13 | 429.82 | 511.04 | 364.29 | 1.44 | 1.47 | 1.42 |
| African Region | 580.59 | 725.70 | 476.44 | 477.36 | 591.81 | 393.29 | 1.22 | 1.23 | 1.21 |
| 2021 |  |  |  |  |  |  |  |  |  |
| Global | 784.53 | 966.02 | 649.26 | 576.46 | 701.84 | 486.27 | 1.36 | 1.38 | 1.34 |
| Eastern Mediterranean Region | 885.62 | 1123.95 | 718.38 | 702.24 | 854.00 | 575.14 | 1.26 | 1.32 | 1.25 |
| European Region | 963.80 | 1202.67 | 792.66 | 657.98 | 800.01 | 548.42 | 1.46 | 1.50 | 1.45 |
| Region of the Americas | 1115.49 | 1407.77 | 909.94 | 802.04 | 987.50 | 660.66 | 1.39 | 1.43 | 1.38 |
| South-East Asia Region | 693.95 | 850.01 | 573.34 | 512.41 | 623.02 | 424.35 | 1.35 | 1.36 | 1.35 |
| Western Pacific Region | 678.82 | 829.63 | 562.44 | 464.04 | 560.07 | 389.17 | 1.46 | 1.48 | 1.45 |
| African Region | 657.70 | 819.67 | 537.75 | 542.65 | 669.98 | 447.05 | 1.21 | 1.22 | 1.20 |

WHO: World Health Organization. The confidence interval is 95%.
Incidence rates: Age-standardized incidence rates.

### Sex difference in incidence of BD and ANX by age

In 2021, the global incidence of BD among individuals under the age of 20 increased rapidly with age for both males and females. The highest incidence rates were 89.77 (95% UI: 65.12 to 121.90) for females and 83.75 (95% UI: 60.97 to 113.38) for males in the age group of 15-19 (Fig. 2A). The absolute sex difference in incidence rates increased rapidly with age for those under 20 years old, with the greatest absolute difference being 6.16 in the age group of 15-19 (Fig. 2B). The relative sex difference in incidence rates increased rapidly with age for those under 20 and over 80 years old, with the greatest relative difference being 1.09 in the age group of 30-34 (Fig. 2C). Supplementary Table 1 shows the incidence of BD in females and males of different ages worldwide in 2021.

**Fig 2.**
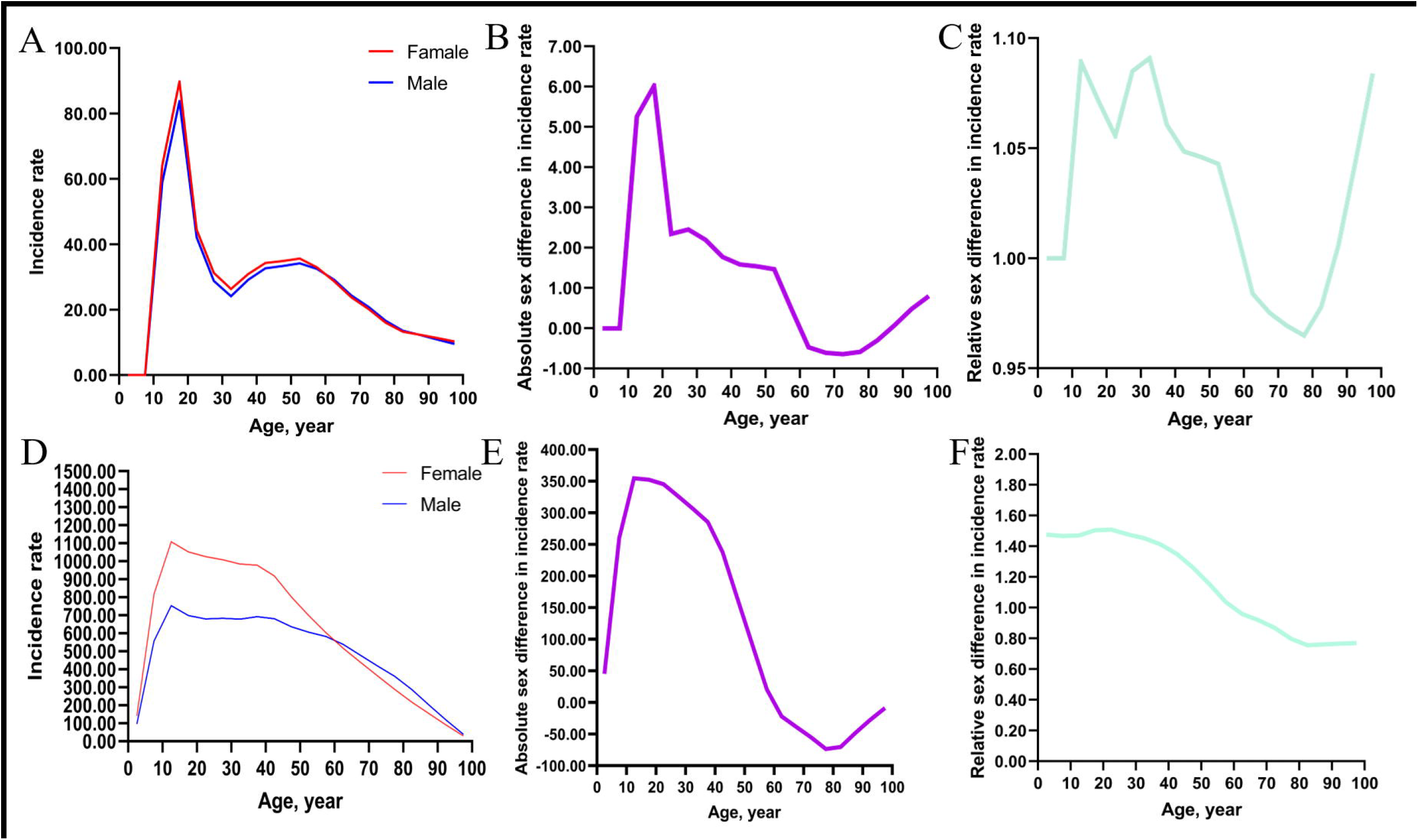
Sex difference in global incidence of BD and ANX in different age groups in 2021. (A) Incidence rates of BD. (B) Absolute (female minus male) sex difference in incidence rates of BD. (C) Relative (female to male ratio) sex difference in incidence rates of BD. (D) Incidence rates of ANX. (E) Absolute sex difference in incidence rates of ANX. (F) Relative sex difference in incidence rates of ANX.

In addition, the global incidence of ANX among individuals under the age of 15 increased rapidly with age for both sexes. The highest incidence rates were 1107.98 (95% UI: 846.66 to 1454.45) for females and 753.22 (95% UI: 569.77 to 998.24) for males in the age group of 10-14 (Fig. 2A). The absolute sex difference in incidence rates increased rapidly with age for those under 20 years old, with the greatest absolute difference being 354.76 in the age group of 10-14 (Fig. 2B). The relative sex difference in incidence rates decreased with age for those over 25 years old, with the greatest relative difference being 1.51 in the age group of 20-24 (Fig. 2C). Supplementary Table 2 shows the incidence of ANX in females and males of different ages worldwide in 2021.

### Sex difference in incidence of BD and ANX by sociodemographic index

ASRs of BD and ANX for males and females in 204 countries and territories in 2021 were demonstrated in Fig. 3A, B, E, F. Absolute and relative sex difference in ASRs of in 204 countries and territories in 2021 were demonstrated in Fig. 3C, D, G, H.

**Fig 3.**
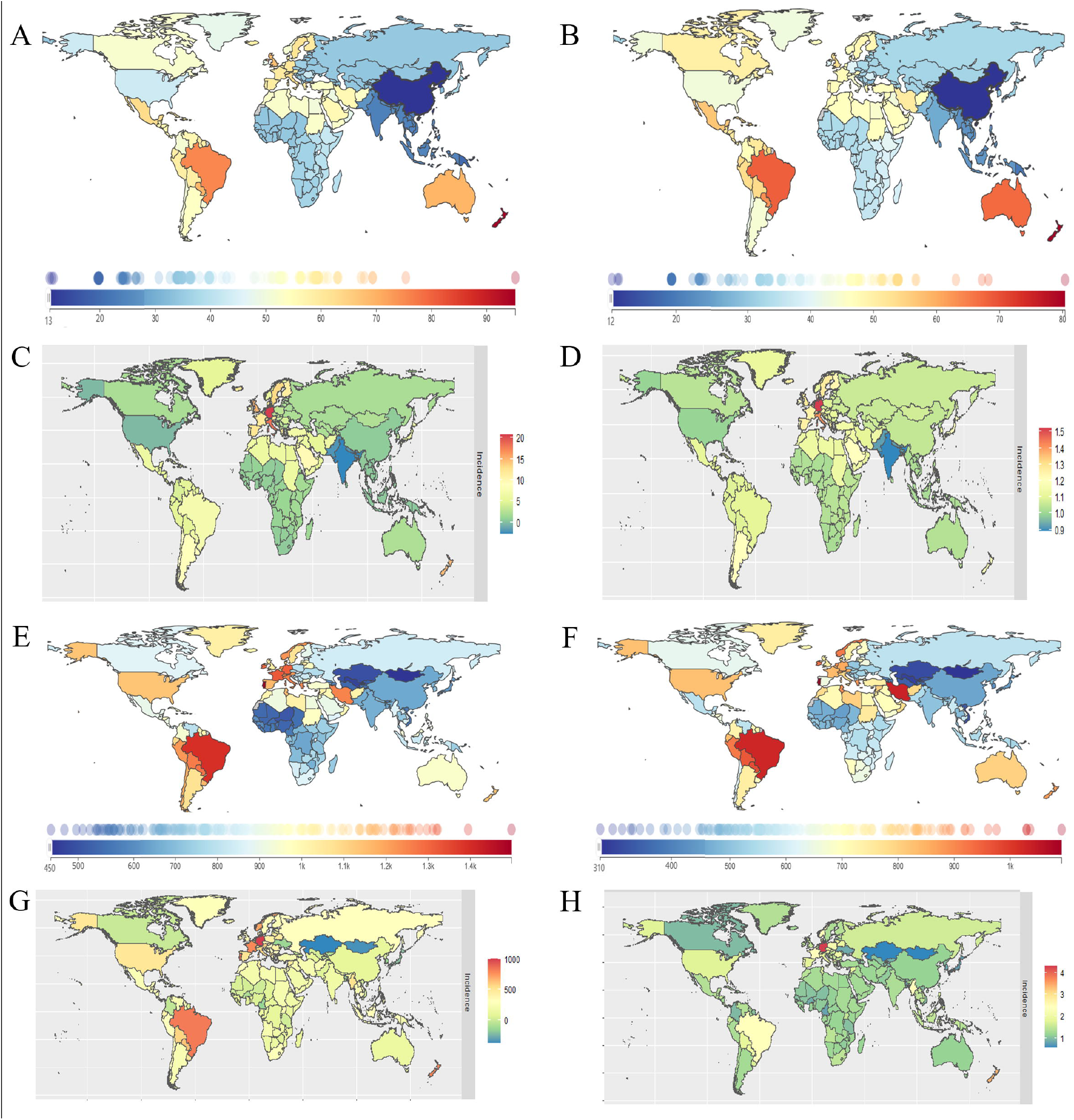
Sex difference in national incidence of BD and ANX in 2021. (A) Age standardized incidence rates of BD in females. (B) Age standardized incidence rates of BD in males. (C)Absolute (female minus male) sex difference in age standardized incidence rates of BD. (D) Relative (female to male ratio) sex difference in age standardized incidence rates of BD. (E) Age standardized incidence rates of ANX in females. (F) Age standardized incidence rates of ANX in males. (G)Absolute (female minus male) sex difference in age standardized incidence rates of ANX. (H) Relative (female to male ratio) sex difference in age standardized incidence rates of ANX.

Mann–Whitney U test showed that females had significant higher ASR of BD than males for 204 countries and territories (Z = −2.04, *P* < 0.001), with median (interquartile range) of ASRs being 39.66 (33.99 to 56.40) for females and 37.62 (31.94 to 47.43) for males. After dividing the groups by SDI, multiple comparisons revealed that females had significantly higher ASR than males in high-middle SDI group (females vs. males: 39.46 (32.28 to 58.99) vs. 33.56 (30.10 to 47.13); Z = −5.91, *P* = 0.039) and high SDI group (54.69 (37..42 to 59.43) vs. 46.43 (32.60 to 47.63); Z = −8.27, *P* < 0.001) (Fig. 4A).

**Fig 4.**
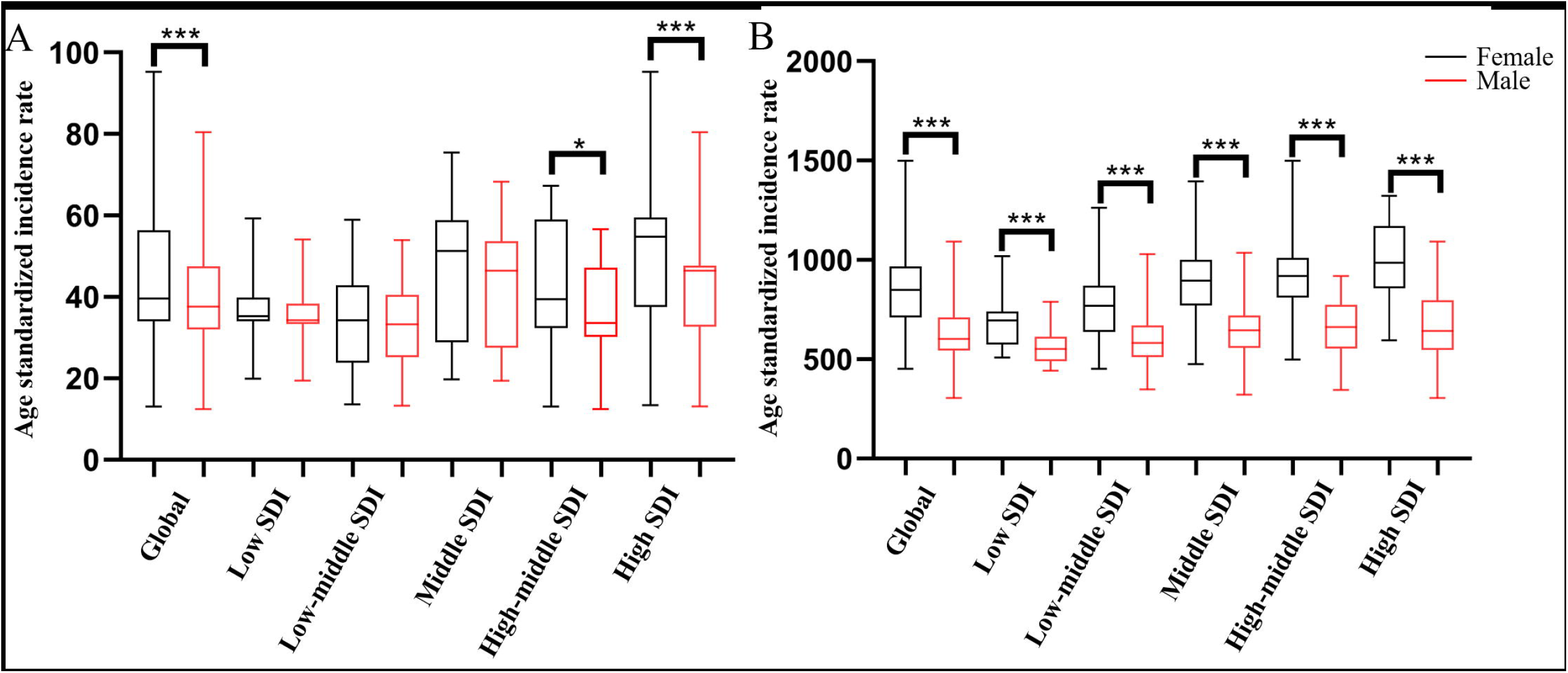
Sex difference in incidence of BD and ANX for SDI-based country groups. (A) Age-standardized incidence rates of BD among females and males for SDI-based country groups. (B) Age-standardized incidence rates of ANX among females and males for SDI-based country groups.Lines inside the boxes indicate the medians, boxes the 25th and 75th percentiles, and lines outside the boxes the minimum and the maximum. ^*^ indicates *P* < 0.05; ^***^ indicates *P* < 0.001. SDI, sociodemographic index.

Additionally, Mann–Whitney U test also showed that he ASR of ANX in females was significantly higher than that in males (Z = −246.26, *P* < 0.001): median (interquartile range) of ASRs was 848.77 (709.81 to 967.93) for females and 602.51 (544.04 to 711.66) for males. Multiple comparisons revealed that females had significantly higher ASR than males, for low SDI group (males vs. females: 694.28 (574.09 to 740.55) vs. 551.46 (488.90–612.80); Z = −142.82, *P* < 0.001), low-middle SDI group (768.53 (637.44 to 868.93) vs. 580.82 (511.16 to 669.80); Z = −187.71, P < 0.001), middle SDI group (895.81 (768.41 to 999.39) vs. 645.37 (555.47 to 719.93); Z = −250.44, P < 0.001), high-middle SDI group (918.97 (810.98 to 1009.92) vs. 660.86 (554.01 to 773.67); Z = −258.11, *P* < 0.001), and high SDI group (984.02 (856.39 to 1170.26) vs. 641.80 (546.04 to 795.65); Z = −342.23, *P* < 0.001) (Fig. 4B).

As shown in Fig. 5, absolute and relative sex differences of BD and ANX were significantly and positively correlated with SDI: Absolute sex differences of BD, standardized β = 12.46 (95% CI, 9.76 to 15.15), *P* < 0.001; Relative sex differences of BD, standardized β = 0.27 (95% CI, 0.22 to 0.33), *P* < 0.001; Absolute sex differences of ANX, standardized β = 444.65 (95% CI, 293.74 to 595.56), *P* < 0.001; Relative sex differences of ANX, standardized β = 0.80 (95% CI, 0.47 to 1.14), *P* < 0.001.

**Fig 5.**
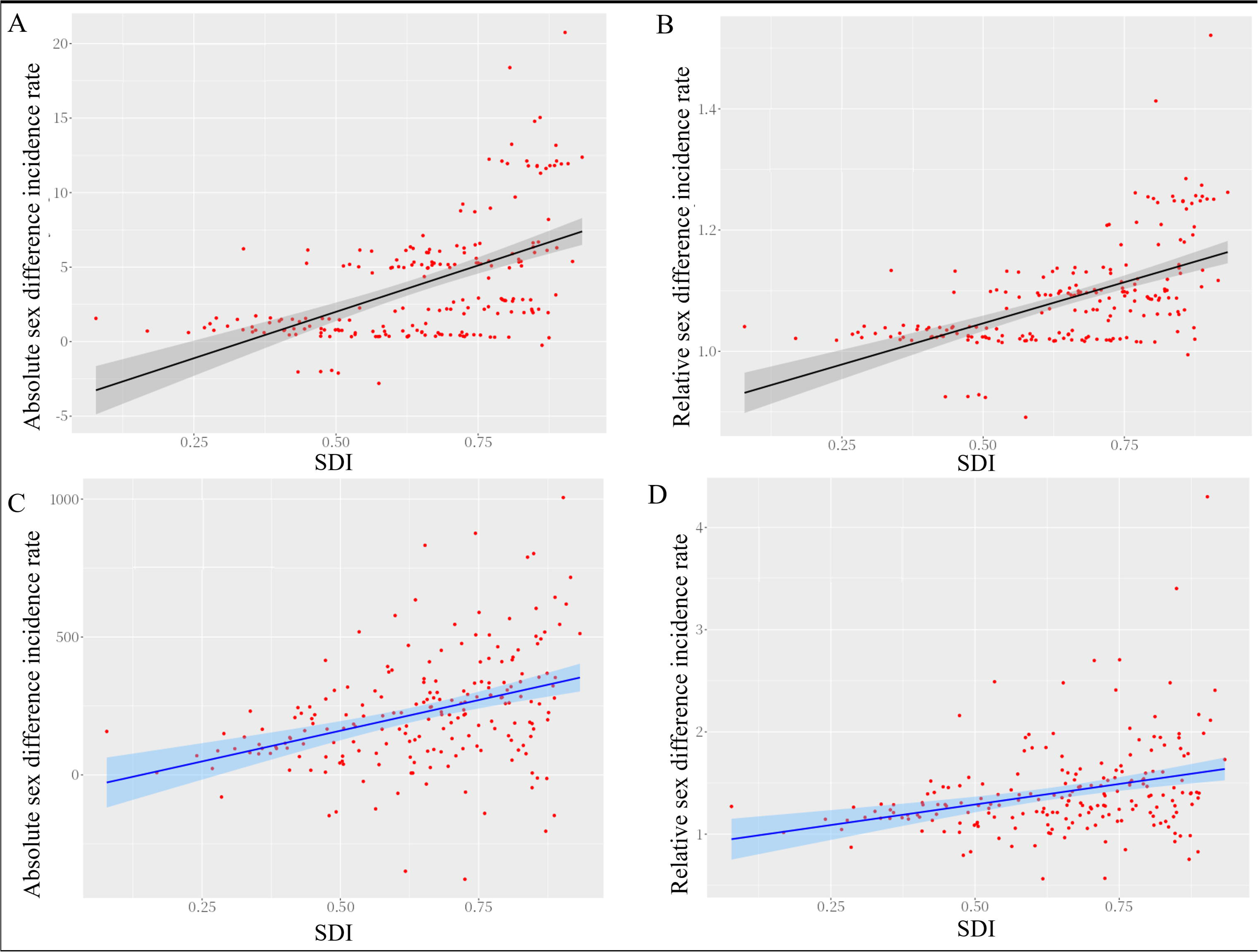
Sex difference in incidence of BD and ANX by SDI. (A) Linear relationship between absolute (female minus male) sex difference in age standardized incidence rates of BD and SDI. (B) Linear relationship between relative (female to male ratio) sex difference in age standardized incidence rates of BD and SDI. (C) Linear relationship between absolute (female minus male) sex difference in age standardized incidence rates of ANX and SDI. (D) Linear relationship between relative (female to male ratio) sex difference in age standardized incidence rates of ANX and SDI. SDI, sociodemographic index.

## Discussion

Sex differences in the incidence of BD and ANX have persisted globally from 1990 to 2019, with rates always higher in women than in men. It is worth noting that the sex difference in BD and ANX incidence increases rapidly with age (under 20 years). In addition, countries with higher levels of socioeconomic development have greater gender differences in BD and ANX incidence.

Firstly, the results of sex difference in incidence by year showed that the incidence was consistently higher in females than in males. Previous studies have shown that the incidence of bipolar disorder and anxiety disorders is lower in men and higher in women [21, 22]. The ASRs for BD in females and males has not changed much globally although ASRs increased globally for both sexes. Over the past 30 years, gender differences in ASRs for bipolar disorder have gradually improved. In contrast, anxiety disorders have demonstrated an increasing trend in incidence and their sex differences. In terms of psychosocial factors, gender socialization puts women in a position of increased mental health vulnerability; women are allowed and encouraged to express their fears and worries, and they are more likely to seek help [23,24]. In contrast to masculine instrumentality or some male characteristics like competitiveness, femininity combined by poor self-esteem increased the development of anxiety [25, 26]. Furthermore, differences between sexes, such as brain regions involved in emotional regulation, sex hormone changes, and genetic factors, all contribute to gender differences in disease [27-29]. More specifically, there was a marked upward trend in the ASR of ANX from 2019 to 2021. This may be related to the COVID-19 pandemic, which has had a more serious impact on women’s mental health than men’s, with higher levels of anxiety [30]. Therefore, the issue of gender differences in BD and the issue of sex differences in ANX remains a major global challenge in the future.

Secondly, sex differences in the incidence of BD and ANX by age revealed that the sex differences in the incidence of BD and ANX increased more significantly in younger age groups. Studies examining mental disease in various age and sex groups have shown that while there is minimal difference in the oldest age group between the sexes, females are more likely than males to experience mental illness in the youngest age group [31]. Sex hormones, the HPA axis, and the immune system interact to alter brain functioning and increase the vulnerability to mood disorders [32]. The hypothalamic-pituitary-gonadal axis stimulates the production of sex hormones differently during adolescence. Additionally, the maturation of the hypothalamic-pituitary-adrenal axis during early adolescence causes a sharp increase in testosterone levels in males and estrogen levels in females [33]. Along with the HPA axis, the immune system undergoes significant development during puberty and is greatly influenced by sex hormones. The immune system is significantly affected by sex and differences in immune responses, and it may be due to the greater concentration of androgen in males than in females, additionally, testosterone not only attenuates the activation of the HPA axis, but it also suppresses the immune response [32]. The increased immune response in females may play a significant role in the vulnerability to mental illnesses [34].

Furthermore, our results indicated that There are regional differences in the incidence of these diseases and the higher the degree of socio-demographic development, the greater the gender differences in the incidence of diseases. In countries with higher living conditions, the negative impact of emotion was greater for women than for men, and women had more mental disease, such as having more symptoms, diagnoses, or feelings of depression and remembering more nightmares [35]. In all WHO regions, especially the regions of Europe and the Americas, the incidence of BD is higher in women than in men (women have about 1.2 times the risk of men), and the incidence of ANX is higher in women than in men (women have about 1.5 times the risk of men). In countries with higher living conditions, psychosocial factors such as gender socialization may have less influence, giving women and men more freedom to pursue the values they care about, leading to greater gender differences [36]. In addition to mental illness, personality, verbal episodic memory, verbal ability, aggressive behavior, and general self-esteem also show greater gender differences in countries with higher living conditions, these factors influence people’s behavior and choices[35]. This gives women and men more freedom to pursue the values they care about, not more of social-role expectations, which also contributes to greater gender disparities [35, 37]. Instead of the traditional gender division of labor based on physical attributes, greater economic prosperity will change the structure of education and careers [38]. With the emergence of new jobs that are considered appropriate for females’ roles in society, more women are being socialized to fit into the careers they pursue, which brings with it mental health problems. Women who worked as housekeepers at home reported less mental health issues than those who worked in countries with southern regimes [11]. According to a representative European study, women who were living alone—especially those who were employed—scored higher for depression [39].

This study is constrained by GBD 2021, including data sources, statistical methods, and so forth [40]. The primary limitation of GBD analysis lies in the availability of raw data. Since country-level aggregated data are employed rather than regional data, potential bias may be introduced in geographic differences of incidence estimates. Although this study offers a global perspective on gender differences in the incidence of BD and ANX, the conclusions may not be applicable to specific regions. Considering that GBD data will be updated in the future, sex differences in the global incidence of BD and ANX over longer time periods can be further explored.

Overall, the findings highlight the importance of tailoring mental health interventions for BD and ANX to address the unique needs of different populations, particularly in regions with pronounced sex differences.

## Conclusion

This study provides a comprehensive analysis of sex differences in the incidence of bipolar disorder (BD) and anxiety disorders (ANX) using data from the Global Burden of Disease Study 2021. Our study revealed that sex difference in the incidence of anxiety disorders and bipolar disorder have persisted worldwide over the past several decades, and the rates have consistently been higher among females than males. The sex difference in the global incidence of bipolar disorder has shown a slight improvement, but that in the global incidence of anxiety disorders has not been effectively mitigated. The sex difference is even more pronounced at younger ages and in more developed nations. These findings underscore the need for continued focus on sex-specific strategies in the prevention, diagnosis, and treatment of bipolar and anxiety disorders.

## Supporting information

Supplementary Contents

## Data Availability

The datasets analyzed during the current study are available from the publicly available website: https://www.healthdata.org/research-analysis/gbd.

## Acknowledgments

We highly praise the collaborators of the 2021 Global Burden of Disease study for their extremely comprehensive analysis of various diseases on a global scale.

## Notes

**Conflicts of interest:** The authors declare that they have no known competing financial interests or personal relationships that could have appeared to influence the work reported in this paper.

### Competing Interest Statement

The authors have declared no competing interest.

### Funding Statement

This study did not receive any funding

### Summary of Updates

Previously, due to the format issues with the uploaded images, they have now been revised to make them correct.

