## Supplementary Contents for "Sex difference in incidence of bipolar and anxiety disorders: findings from the Global Burden of Disease Study 2021"

**Table S1**. The global age-specific incidence of bipolar disorder for both sexes in 2021.

| Age (years) | Female |  |  |  | Male |  |  |  | Risk ratio | | |
| --- | --- | --- | --- | --- | --- | --- | --- | --- | --- | --- | --- |
|  | Incidence rates | Upper | Lower |  | Incidence rates | Upper | Lower |  | Mean | Upper | Lower |
| All ages | 33.23 | 39.59 | 27.88 |  | 31.76 | 37.80 | 26.64 |  | 1.05 | 1.05 | 1.05 |
| <5 | 0.00 | 0.00 | 0.00 |  | 0.00 | 0.00 | 0.00 |  | None | None | None |
| 5-9 | 0.00 | 0.00 | 0.00 |  | 0.00 | 0.00 | 0.00 |  | None | None | None |
| 10-14 | 64.16 | 91.64 | 43.42 |  | 58.91 | 83.43 | 40.26 |  | 1.09 | 1.10 | 1.08 |
| 15-19 | 89.77 | 121.90 | 65.12 |  | 83.75 | 113.38 | 60.97 |  | 1.07 | 1.08 | 1.07 |
| 20-24 | 44.49 | 76.53 | 21.12 |  | 42.15 | 72.03 | 20.13 |  | 1.06 | 1.06 | 1.05 |
| 25-29 | 31.28 | 50.23 | 16.63 |  | 28.83 | 46.85 | 15.33 |  | 1.09 | 1.09 | 1.07 |
| 30-34 | 26.37 | 44.91 | 14.06 |  | 24.18 | 41.52 | 12.83 |  | 1.09 | 1.10 | 1.08 |
| 35-39 | 30.93 | 50.01 | 14.81 |  | 29.16 | 47.10 | 14.17 |  | 1.06 | 1.06 | 1.05 |
| 40-44 | 34.30 | 56.15 | 16.96 |  | 32.71 | 52.55 | 16.45 |  | 1.05 | 1.07 | 1.03 |
| 45-49 | 34.90 | 49.60 | 21.64 |  | 33.36 | 47.29 | 20.81 |  | 1.05 | 1.05 | 1.04 |
| 50-54 | 35.67 | 57.66 | 18.44 |  | 34.20 | 55.39 | 17.80 |  | 1.04 | 1.04 | 1.04 |
| 55-59 | 32.96 | 48.57 | 18.76 |  | 32.48 | 47.37 | 18.87 |  | 1.01 | 1.03 | 0.99 |
| 60-64 | 28.71 | 46.14 | 14.55 |  | 29.18 | 46.10 | 15.30 |  | 0.98 | 1.00 | 0.95 |
| 65-69 | 23.80 | 36.59 | 14.98 |  | 24.41 | 37.06 | 15.57 |  | 0.98 | 0.99 | 0.96 |
| 70-74 | 20.26 | 28.83 | 13.75 |  | 20.90 | 29.36 | 14.54 |  | 0.97 | 0.98 | 0.95 |
| 75-79 | 16.04 | 25.29 | 9.77 |  | 16.63 | 25.82 | 10.24 |  | 0.96 | 0.98 | 0.95 |
| 80-84 | 13.31 | 23.04 | 6.48 |  | 13.61 | 23.08 | 6.81 |  | 0.98 | 1.00 | 0.95 |
| 85-89 | 12.34 | 19.21 | 7.29 |  | 12.27 | 19.08 | 7.24 |  | 1.01 | 1.01 | 1.01 |
| 90-94 | 11.37 | 17.77 | 6.57 |  | 10.89 | 17.21 | 6.22 |  | 1.04 | 1.06 | 1.03 |
| 95 plus | 10.33 | 19.97 | 4.25 |  | 9.53 | 18.66 | 3.92 |  | 1.08 | 1.08 | 1.07 |

The confidence interval is 95%.

**Table S2**. The global age-specific incidence of anxiety disorder for both sexes in 2021.

| Age (years) | Female |  |  |  | Male |  |  |  | Risk ratio | | |
| --- | --- | --- | --- | --- | --- | --- | --- | --- | --- | --- | --- |
|  | Incidence rates | Upper | Lower |  | Incidence rates | Upper | Lower |  | Mean | Upper | Lower |
| All ages | 781.54 | 960.74 | 646.66 |  | 585.63 | 710.05 | 491.94 |  | 1.33 | 1.35 | 1.31 |
| <5 years | 141.1 | 204.41 | 94.31 |  | 95.54 | 137.4 | 63.06 |  | 1.48 | 1.49 | 1.48 |
| 5-9 years | 817.76 | 1170.15 | 551.33 |  | 557.35 | 796.93 | 370.81 |  | 1.47 | 1.49 | 1.47 |
| 10-14 years | 1107.98 | 1454.45 | 846.66 |  | 753.22 | 998.24 | 569.77 |  | 1.47 | 1.49 | 1.46 |
| 15-19 years | 1051.35 | 1488.87 | 665.04 |  | 698.61 | 965.98 | 449.53 |  | 1.5 | 1.54 | 1.48 |
| 20-24 years | 1025.17 | 1502.34 | 582 |  | 679.71 | 981.44 | 388.06 |  | 1.51 | 1.53 | 1.5 |
| 25-29 years | 1008.71 | 1375.47 | 722.23 |  | 682.58 | 915.31 | 496.56 |  | 1.48 | 1.5 | 1.45 |
| 30-34 years | 984.1 | 1290.47 | 724.55 |  | 677.53 | 876.38 | 497.74 |  | 1.45 | 1.47 | 1.45 |
| 35-39 years | 977.71 | 1349.54 | 664.02 |  | 692.11 | 948.73 | 464.19 |  | 1.41 | 1.42 | 1.41 |
| 40-44 years | 918.18 | 1289.68 | 595.92 |  | 680.2 | 946.23 | 451.33 |  | 1.35 | 1.36 | 1.32 |
| 45-49 years | 800.89 | 1083.24 | 568.83 |  | 635.98 | 833.7 | 469.57 |  | 1.26 | 1.3 | 1.21 |
| 50-54 years | 698.16 | 924.58 | 511.75 |  | 605.54 | 796.84 | 459.77 |  | 1.15 | 1.16 | 1.11 |
| 55-59 years | 602.71 | 855.88 | 411.95 |  | 581.92 | 822.67 | 401.28 |  | 1.04 | 1.04 | 1.03 |
| 60-64 years | 516.56 | 763.9 | 328.45 |  | 538.42 | 767.18 | 367.93 |  | 0.96 | 1 | 0.89 |
| 65-69 years | 439.38 | 618.88 | 307.8 |  | 477.88 | 647.2 | 352 |  | 0.92 | 0.96 | 0.87 |
| 70-74 years | 363.19 | 493.36 | 271.8 |  | 418.26 | 550.16 | 319.4 |  | 0.87 | 0.9 | 0.85 |
| 75-79 years | 287.65 | 401.12 | 201.17 |  | 361.01 | 486.75 | 258.33 |  | 0.8 | 0.82 | 0.78 |
| All ages | 781.54 | 960.74 | 646.66 |  | 585.63 | 710.05 | 491.94 |  | 1.33 | 1.35 | 1.31 |
| 80-84 years | 216.68 | 332.08 | 133.53 |  | 286.95 | 418.21 | 185.22 |  | 0.76 | 0.79 | 0.72 |
| 85-89 years | 152.72 | 234.22 | 94 |  | 200.69 | 293.12 | 129.61 |  | 0.76 | 0.8 | 0.73 |
| 90-94 years | 89.63 | 137.37 | 55.24 |  | 116.89 | 171.04 | 75.53 |  | 0.77 | 0.8 | 0.73 |
| 95+ years | 29.05 | 44.46 | 17.95 |  | 37.73 | 55.26 | 24.49 |  | 0.77 | 0.8 | 0.73 |

The confidence interval is 95%.
